# Estimation of free water-corrected microscopic fractional anisotropy

**DOI:** 10.1101/2022.10.27.22281560

**Authors:** Nico J. J. Arezza, Mohammad Omer, Corey A. Baron

**Author notes:** **Correspondence:** Nico Arezza.

## Abstract

Water diffusion anisotropy MRI is sensitive to microstructural changes in the brain that are hallmarks of various neurological conditions. However, conventional metrics like fractional anisotropy are confounded by neuron fiber orientation dispersion, and the relatively low resolution of diffusion-weighted MRI gives rise to significant free water partial volume effects in many brain regions. Microscopic fractional anisotropy is a recent metric that can report water diffusion anisotropy independent of neuron fiber orientation dispersion but is still susceptible to free water contamination. In this paper, we present a free water elimination (FWE) technique to estimate microscopic fractional anisotropy and other related diffusion indices by implementing a model in which the MRI signal within a voxel is assumed to come from two distinct sources: a tissue compartment and a free water compartment. A two-part algorithm is proposed to rapidly fit a set of diffusion-weighted MRI volumes containing both linear- and spherical-tensor encoding acquisitions to the model. Simulations and *in vivo* acquisitions with four healthy volunteers indicated that the FWE method may be a feasible technique for measuring microscopic fractional anisotropy and other indices with greater specificity to neural tissue characteristics than conventional methods.

## Introduction

Diffusion-weighted MRI (dMRI) is a non-invasive imaging modality that uses specialized pulse sequences to sensitize the MRI signal to the random molecular motion of water (Stejskal and Tanner 1965; Tanner 1965). On MRI-relevant time frames, water molecules traverse microscopic length scales in tissue, and their diffusion is dictated by the presence of restricting boundaries such as cell membranes and other structures. By exploiting the known relationships between dMRI signal and tissue properties, dMRI measurements can act as surrogate indicators of physical properties of neural tissue, and this capability has led to dMRI finding use in the study of neurological disorders like multiple sclerosis (Inglese and Bester 2010; Rovaris et al. 2005), Alzheimer’s disease (Yu Zhang et al. 2009), and stroke (van Everdingen et al. 1998), among others.

The most widely used dMRI technique is diffusion tensor imaging (DTI), in which dMRI data is fitted to the diffusion tensor model. DTI is based on the first order cumulant expansion of the logarithm of the dMRI signal as a function of diffusion weighting or b-value (Basser, Mattiello, and LeBihan 1994; Frisken 2001), which can be represented by the equation:

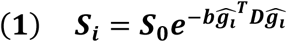

where *S*_*i*_ is the diffusion-weighted MRI signal of a particular acquisition *i, S*_0_ is the MRI signal in the absence of diffusion weighting, *b* is the b-value, which describes the strength of the diffusion weighting, ***D*** is the diffusion tensor, and 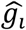 is a unit vector describing the direction of the diffusion-sensitizing gradient. DTI requires linear tensor encoding (LTE) acquisitions in different diffusion directions at a single b-value plus one or more acquisitions with no diffusion weighting and can report metrics such as the mean diffusivity (*D*) and fractional anisotropy (FA) of water diffusion. However, the DTI model assumes that diffusion follows a mono-Gaussian distribution, which is a reasonable assumption only at low b-values (Johansen-Berg and Behrens 2009). The diffusion kurtosis imaging (DKI) model further expands the cumulant expansion of the logarithm of the dMRI signal to the second order to account for non-Gaussian diffusion, but requires the acquisition of dMRI data at two or more b-values (Jensen et al. 2005). The “mean signal” DKI model, in which data acquired from all diffusion directions are averaged into a single image volume known as the powder average, can be represented as:

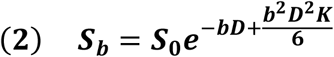

where *S*_*b*_ is the dMRI signal of the powder averaged data and *K* is the diffusion kurtosis (Jensen et al. 2005). All models presented henceforth in this article are similarly based on powder averaged data, fitting to scalar variables such as *D* in place of tensors such as ***D***, though it should be noted that the tensor-based models are required to estimate FA.

The DTI and DKI models are limited by two major factors that affect their specificity to neuronal microstructure: (1) the tensor models used to measure anisotropy are sensitive to neuron fiber orientation, causing FA to be underestimated in brain regions containing crossing or fanning axons (Jones, Knösche, and Turner 2013; Szczepankiewicz et al. 2015), and (2) the presence of extracellular free water within the voxel of interest biases diffusion measurements in both the tensor and mean signal models, potentially confounding or masking true microstructural changes within the tissue (Jones and Cercignani 2010; Vos et al. 2011; Alexander et al. 2001; C. A. Baron and Beaulieu 2015). Typically, a voxel with free water contamination will have elevated *D* and reduced FA due to the high diffusivity and negligible anisotropy of free water.

To overcome the first limitation, techniques such as microscopic fractional anisotropy (μFA) imaging, which reports water diffusion anisotropy independent of the neuron fiber orientation dispersion, were developed (Jespersen et al. 2013; Lasič et al. 2014; Shemesh et al. 2016). μFA can be estimated by fitting traditional LTE dMRI data to various constrained models using a priori knowledge of the underlying tissue (Novikov et al. 2019; Kaden, Kruggel, and Alexander 2016; Kaden et al. 2016), but more accurate measures of μFA can be acquired using advanced dMRI sequences like double diffusion encoding (Cory, Garroway, and Miller 1990; Henriques, Jespersen, and Shemesh 2020) or spherical tensor encoding (STE) (Lasič et al. 2014; Szczepankiewicz et al. 2015; Westin et al. 2016). Previous studies have demonstrated that μFA may be more suitable than FA for a number of applications such as in evaluating white matter degeneration in Parkinson’s disease (Ikenouchi et al. 2020), delineating lesions in multiple sclerosis (Yang et al. 2018), and differentiating between different types of brain tumors (Szczepankiewicz et al. 2015).

The bias caused by free water contamination on DTI and DKI measurements results from the fact that indices quantified using both models represent the weighted average of all diffusion compartments within a voxel rather than markers of a specific tissue. The diffusivity of free water at 37°C is isotropic and approximately 3-4 times higher than that of brain tissue, so it has a significant effect on the voxel-level dMRI parameters, even at low volume fractions (Pierpaoli and Basser 1996). Moreover, the free water signal is typically a factor of 2-3 times higher than brain-tissue for the T^2^-weighted scans used for dMRI, which further exacerbates these partial volume effects. Accordingly, dMRI measurements made in brain regions with significant free water partial volumes, such as the fornix and ventricle-adjacent regions, are greatly affected (Li et al. 2013; Metzler-Baddeley et al. 2012).

The effects of free water partial volumes can be attenuated by using non-zero minimum diffusion weighting (C. A. Baron and Beaulieu 2015) and by implementing fluid-attenuated inversion recovery dMRI sequences (Papadakis et al. 2002; Chou et al. 2005), but both techniques decrease signal-to-noise ratio (SNR), the former affects DTI metrics in tissue with no free water, and the latter increases specific absorption rate and scan time (Pasternak et al. 2009). Alternatively, modifications to the DTI and DKI models can be used to distinguish between dMRI signal from free water and dMRI signal from functional brain tissue. The free water elimination DTI (FWE-DTI) model separates the dMRI signal into two compartments: one representing the free water compartment and one representing functional brain tissue (Pasternak et al. 2009), and can be expressed as:

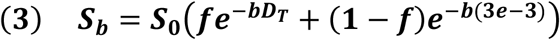

where *f* is the volume fraction of tissue within the voxel of interest and *D*_*T*_ is the mean diffusivity of the tissue compartment. The *(3e-3)* term represents the diffusivity of free water at 37°C in mm^2^/s. This model enables more accurate estimation of tissue-specific indices than traditional DTI and has attracted interest for use in studying neurodegeneration in Alzheimer’s disease (Hoy et al. 2017), Parkinson’s disease (Planetta et al. 2016), and traumatic brain injury (Pasternak, Koerte, et al. 2014). Additionally, the volume fraction metric is a potential surrogate marker for edema (Pasternak et al. 2009; Pasternak, Maier-Hein, et al. 2014). While traditional DTI can be performed using single b-shell data, FWE-DTI should be performed with data collected at multiple b-values to reduce model fitting degeneracies at the expense of increased scan time (Golub, Neto Henriques, and Gouveia Nunes 2020). Recently, the FWE-DTI model was expanded to incorporate the kurtosis term to account for non-Gaussian diffusion in the tissue compartment; this modification to FWE-DTI is referred to as the free water elimination DKI (FWE-DKI) model (Collier et al. 2018):

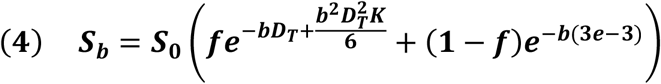

In this article, we propose a technique to measure water diffusion anisotropy that combines a common μFA acquisition protocol to achieve insensitivity to neurite orientation with the free water elimination models’ ability to distinguish between free water partial volume effects and true tissue properties. Previously, we demonstrated that μFA can be estimated by jointly fitting multi-shell LTE and STE dMRI data to the DKI model, as per the following equations (Arezza, Tse, and Baron 2021):

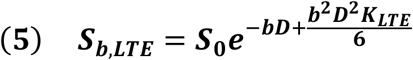

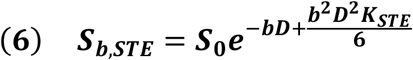

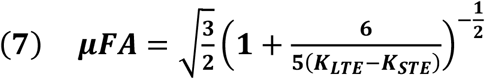

where the subscripts *LTE* and *STE* denote the encoding scheme. By combining equations (5) and (6) with a FWE model, a joint fit of LTE and STE data to the FWE-DKI model can be performed:

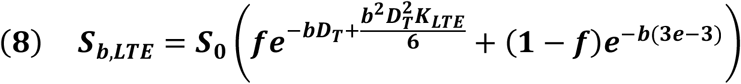

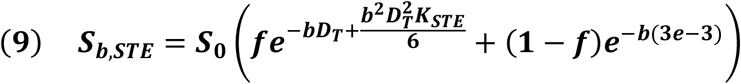

The *D* and *K* terms obtained using equations (8) and (9) characterize water diffusion in brain tissue independent of free water. Accordingly, μFA estimated from equation (7) using these corrected indices should characterize water diffusion anisotropy in tissue free of the bias caused by free water partial volumes. This imaging strategy which combines the FWE-DKI model with μFA imaging acquisition will be referred to herein as the FWE imaging method, whereas the technique that involves fitting the data to the DKI model will be referred to as the conventional method.

## Materials and Methods

### Fitting Algorithm

In this work, a two-part algorithm (denoted Part I and Part II) was used to obtain a solution to the joint fitting of STE and LTE data. In the first part of the algorithm, low b-value (b≤1000s/mm^2^) powder average STE data were fitted to the FWE-DTI model to obtain estimates of volume fraction and mean tissue diffusivity. Those indices were used as initial guesses in the second part of the algorithm, in which powder average STE and powder average LTE data across all b-values were jointly fitted to equations (8) and (9).

Part I of the algorithm exploits the FWE-DTI model’s lower complexity relative to the FWE-DKI model, reducing the number of unknown variables to be solved for by omitting the kurtosis term. The effects of non-Gaussian diffusion on dMRI, while deleterious to DTI and FWE-DTI fits, are minimal at low b-values; thus, *f* and *D*_*T*_ can be initially estimated despite omitting the second order term in the dMRI signal cumulant expansion. Using only the STE data as input further reduces the effects of non-Gaussian diffusion on the fit because it typically has minimal kurtosis. More specifically, LTE introduces a variance to the powder average signal due to the different diffusion encoding directions used for each acquisition; STE signals are free of this variance and deviate less from the mono-Gaussian diffusion assumption inherent to the FWE-DTI model in tissue-containing voxels (Lasič et al. 2014; Henriques, Jespersen, and Shemesh 2020). In this work, an iterative method was used to solve the FWE-DTI equation. In each iteration, the low b-value STE data were first fitted to the FWE-DTI model (equation (3)) using the least squares method with a fixed estimate of *D*_*T*_ = 7*e* − 4 *mm*^2^*/s* used as an initial guess in the first iteration. Then, a correction was implemented to constrain *f* and (1 − *f*) to be positive. The *D*_*T*_ estimate was then updated by again fitting the data to equation (3) using the least squares method, this time with *f* and (1 − *f*) as fixed inputs. A correction was implemented at the end of each iteration to set *D*_*T*_ to 0 in voxels with very small tissue compartments (*f* < 0.1).

In Part II of the algorithm, the LTE and STE data across all b-values were jointly fitted to the FWE-DKI model using the *f* and *D*_*T*_ indices from Part I as initial estimates. Again, an iterative method was employed that was similar to that of Part I. In each iteration, the data were first fitted to the FWE-DKI model (equations (8) and (9)) to solve for *D*_*T*_, *K*_*LTE*_ and *K*_*STE*_ using a fixed *f* value (the first iteration used the value of *f* that was obtained from Part 1). Corrections were performed to constrain *K*_*LTE*_ to be positive and *K*_*STE*_ to be greater than or equal to -0.1. Then, the data were jointly fitted to equations (8) and (9), this time using fixed estimates of *D*_*T*_, *K*_*LTE*_ and *K*_*STE*_ to obtain an updated estimate of *f*. A final correction was performed at the end of each iteration to constrain *f* and (1 − *f*) to be positive.

Part I and Part II were each performed for 100 iterations for all simulated and *in vivo* implementations of FWE-μFA investigated in this article. For all cases, adding more iterations caused negligible changes in the output parameters. The fitting code is openly available at gitlab.com/coreybaron/fwe_ufa.

### Simulations

To validate the FWE-μFA method proposed herein, equations (8) and (9) were used to generate synthetic LTE and STE powder average signals to simulate white matter (WM) and gray matter (GM) voxels. For each voxel, signals were generated for b-values of 0, 700, 1000, 1400, and 2000 s/mm^2^. To simulate a typical WM configuration, μFA was measured from publicly available dMRI data (C. Baron and Arezza 2020) using the conventional μFA method (Arezza, Tse, and Baron 2021), and typical parameter values were extracted from frontal WM voxels in which free water contamination is expected to be minimal. The corresponding parameters are *D*_*T*_ = 8*e* − 4*mm*^2^*/s, K*_*LTE*_ = 1.2, and *K*_*STE*_ = 0.1, which corresponds to a μFA of 0.85 as per equation (7). These parameters were used to simulate the signal acquired in voxels with tissue volume fractions of 0.25, 0.5, 0.75, and 1 via equations (8) and (9). Rician noise was simulated by adding random Gaussian noise to the signal and then taking the magnitude of the noisy signal. The noise standard deviation was chosen to achieve an SNR of 20 for each non-diffusion weighted acquisition. The standard deviation of the noise added to the signals was scaled by 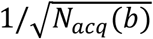, where *N*_*acq*_(*b*) is the number of acquisitions used experimentally (refer to Materials and Methods: *In Vivo*) for each b-value, to account for averaging from multiple acquisitions when the powder average is computed. Notably, PCA denoising (Veraart et al. 2016) is typically used for *in vivo* data prior to parameter fitting and, accordingly, the simulations likely explore a more challenging fitting scenario than *in vivo*.

Due to the presence of free water in cortical GM voxels, as well as the heterogeneity between different deep GM regions of the brain, a typical GM configuration is difficult to assess. For this work, GM μFA was set to 0.55 as this is within the range of values found in the hippocampus (Yoo et al. 2021) and other deep GM regions (Lawrenz, Brassen, and Finsterbusch 2016); using the same *D*_*T*_ as the WM simulations, the *K*_*LTE*_ and *K*_*STE*_ values were set to 0.9 and 0.6, which yields the desired μFA = 0.55 via equation (7). GM simulations were performed over the same tissue volume fractions and SNR as the WM simulations.

1000 realizations of random noise were simulated at each tissue volume fraction for the WM and GM configurations. The FWE and conventional methods of estimating μFA were performed on the simulated voxels, and the mean and standard deviation of the following indices were computed across all setups for both methods: *D*_*T*_, anisotropic kurtosis (*K*_*aniso*_), isotropic kurtosis (*K*_*iso*_), and μFA. The kurtosis terms were computed as follows:

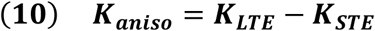

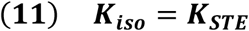

### In Vivo

To assess the FWE μFA algorithm in real dMRI data, 4 healthy volunteers (2 female and 2 male, mean age 28.0 ± 6.6 years) were scanned on a 3T Prisma whole body MRI system (Siemens Healthineers) located in the Centre for Functional and Metabolic Mapping at Western University with 80 mT/m strength and 200 T/m/s slew rate. Volunteers first underwent T1-weighted MPRAGE acquisitions with 1 mm isotropic resolution to provide structural image volumes for segmenting regions-of-interest (ROIs). Then each subject underwent dMRI scans consisting of 5 acquisitions with no diffusion-weighting (b = 0 s/mm^2^), and 3, 15, 6, and 22 LTE acquisitions plus 6, 10, 10, and 27 STE acquisitions at b-values of 700, 1000, 1400, and 2000 s/mm^2^, respectively. The STE pulse sequence used is described in Arezza et al (Arezza, Tse, and Baron 2021). The other parameters for the dMRI acquisitions were: TE/TR = 94/4500 ms, field-of-view = 220 × 200 mm^2^, resolution = 2 mm (isotropic), 48 slices, and rate 2 in-plane parallel imaging combined with rate 2 simultaneous multislice. Note that the b-values acquired in the dMRI acquisitions match those of the simulations.

Post-processing for the dMRI data included PCA denoising (Veraart et al. 2016) and Gibbs ringing correction using MRtrix3 (Kellner et al. 2016; Tournier et al. 2019), and eddy current artifact correction using FSL Eddy (Andersson and Sotiropoulos 2016). Powder average signals were then computed from the LTE and STE data at each b-value and were then fitted to equations (8) and (9) to obtain μFA via the FWE method and fitted to equations (5) and (6) using ordinary least squares to obtain μFA via the conventional method.

The T1-weighted image volumes were used to obtain masks for ROIs because of their superior resolution and soft-tissue contrast compared to the dMRI image volumes. WM ROI masks were generated using the FAST tool from FSL (Y. Zhang, Brady, and Smith 2001) using a probability threshold of 99% and limiting the masks to the region of the brain superior to the thalamus. Masks for the hippocampus, putamen, and thalamus were generated using the FIRST tool from FSL (Patenaude et al. 2011). ROI masks for the fornix were manually drawn. The T1 volumes were then registered to the powder averaged b=0 s/mm^2^ volumes using symmetric diffeomorphic and affine transformations with ANTS software (https://github.com/ANTsX/ANTs); these transformations were then applied to each of the ROI masks to register them to dMRI space.

The ROIs were selected to test several specific hypotheses. The WM and putamen are minimally contaminated by free cerebrospinal fluid, so it was expected that measurements made with the FWE and kurtosis μFA methods would be similar. The thalamus and hippocampus ROIs represent deep GM structures adjacent to free water, in which it was expected that the *D*_*T*_ would be reduced and μFA would be elevated when using the FWE technique due to mitigation of free water signal. The fornix, which is both adjacent to the lateral ventricles and small relative to the image resolution, represents an ROI that is likely to have significant free water contamination; thus, much lower *D*_*T*_ and much higher μFA were expected in this region when the FWE technique was used.

Mean and standard deviation of the following indices were computed in each of the ROIs to compare the FWE and conventional μFA techniques: *D*_*T*_, *K*_*aniso*_, *K*_*iso*_, and μFA. Voxels with *f*<0.25 after fitting were excluded from this analysis because there is very little tissue signal for which the diffusion parameters correspond to, which leads to unstable estimations of the parameters.

## Results

### Simulations

For all volume fractions except *f*=1, the FWE μFA method yielded more accurate measurements of *D*_*T*_ and μFA than the conventional method in both the WM (Figure 1) and GM (Figure 2) configurations. At *f*=0.25, the FWE method overestimated volume fraction in both the WM and GM simulations, with mean estimates of 0.34 and 0.35, respectively; however, resulting *D*_*T*_ and μFA estimates were closer to the ground truth than measurements produced by the conventional method. FWE estimates of *K*_*aniso*_ were higher than estimates produced by the conventional method across all volume fractions, while estimates of *K*_*iso*_ were lower. The variance of parameter estimations over the 1000 repetitions increased for decreasing *f*.

**Figure 1.**
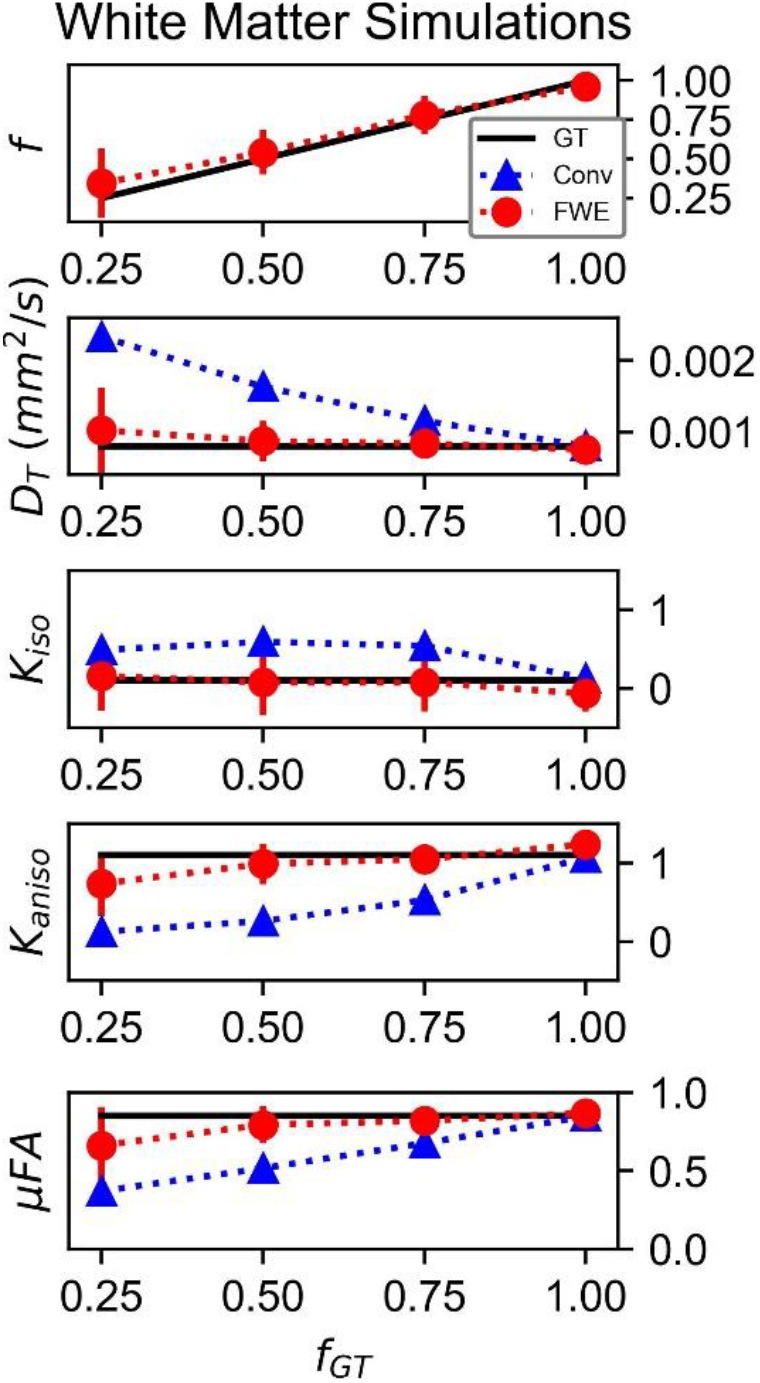
Diffusion MRI indices measured in simulated white matter voxels for various ground truth volume fractions (*f*_*GT*_). The solid black line indicates the ground truth (GT) metric, the blue triangles indicate the mean measurements produced by the conventional (Conv) method, and the red circles indicate the mean measurements produced by the free water elimination (FWE) method. Note that *D*_*T*_ is used interchangeably with *D* for the conventional method.

**Figure 2.**
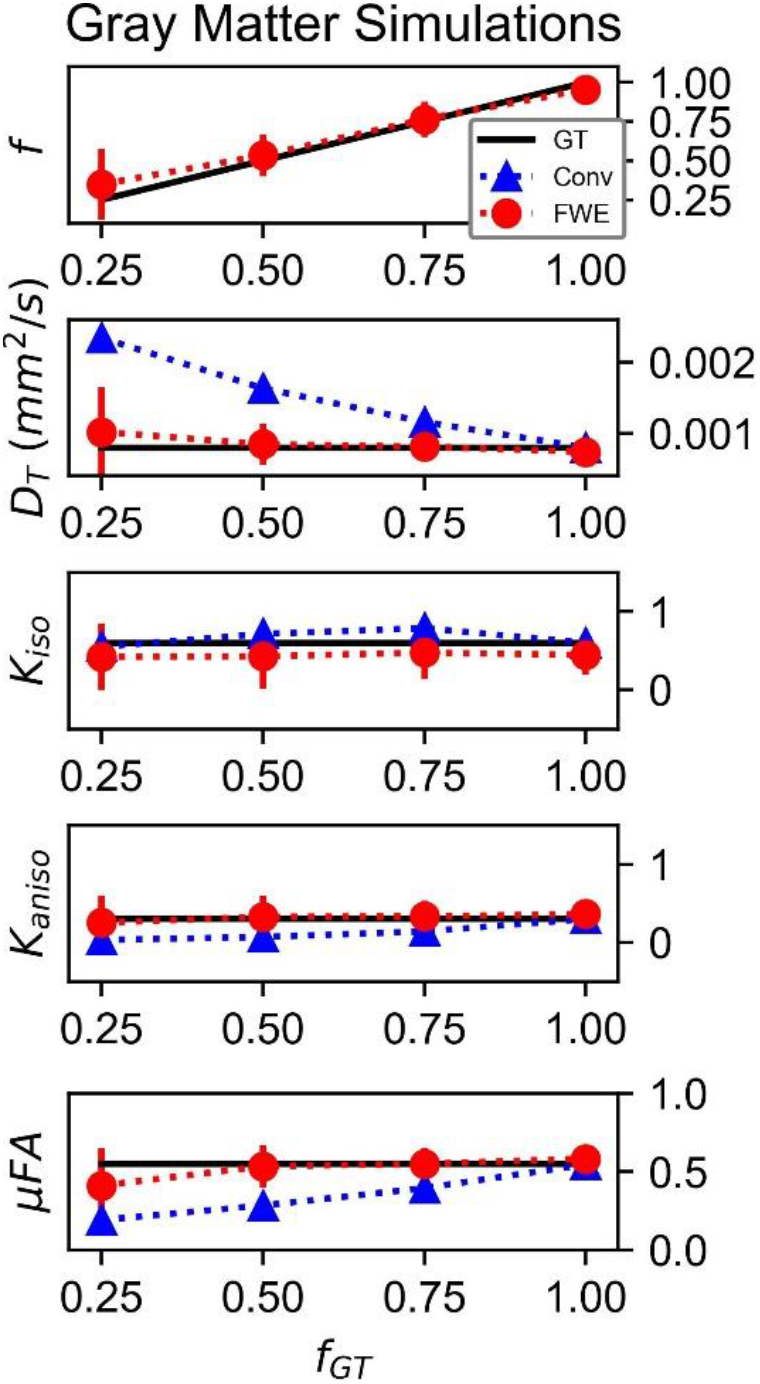
Diffusion MRI indices measured in simulated gray matter voxels for various ground truth volume fractions (*f*_*GT*_). The solid black line indicates the ground truth (GT) metric, the blue triangles indicate the mean measurements produced by the conventional (Conv) method, and the red circles indicate the mean measurements produced by the free water elimination (FWE) method. Note that *D*_*T*_ is used interchangeably with *D* for the conventional method.

### In Vivo

Example slices of *D*_*T*_, *K*_*aniso*_, *K*_*iso*_, and μFA generated with the FWE and conventional methods are depicted in Figure 3, as well as a sample slice depicting voxels with *f* < 0.25. Zoom-ins of a cortical region are depicted in Figure 4, where decreases in *K*_*iso*_ and *D*_*T*_, and increases in μFA and *K*_*aniso*_, are observed for FWE relative to the conventional method throughout the cortex, which agrees with expectations from the simulations. The ROIs are depicted in T1-weighted images in Figure 5, as well as the mean and standard deviations of relevant diffusion indices generated using the two methods. Mean volume fractions in the WM, putamen, hippocampus, thalamus, and fornix regions were 0.96, 0.96, 0.82, 0.82, and 0.64, respectively. Differences in *D*_*T*_ and μFA between the two methods were smallest in the WM and putamen ROIs. When the FWE method was employed, *D*_*T*_ was reduced by 6.4% and 7.5% in the WM and putamen, respectively, compared to measures made using the conventional method, while μFA was increased by 3.5% and 5.3%. Greater differences between methods were observed in the deep GM regions: *D*_*T*_ was reduced by 37.1% in the hippocampus and 42.8% in the thalamus when FWE was used, while μFA was increased by 22.0% and 16.8% in those regions. The most significant differences between methods were observed in the fornix, in which *D*_*T*_ was reduced by 59.2% and μFA was increased by 30.5% when FWE was applied. In all ROIs, mean *K*_*aniso*_ was reduced while mean *K*_*iso*_ was increased when FWE was used.

**Figure 3.**
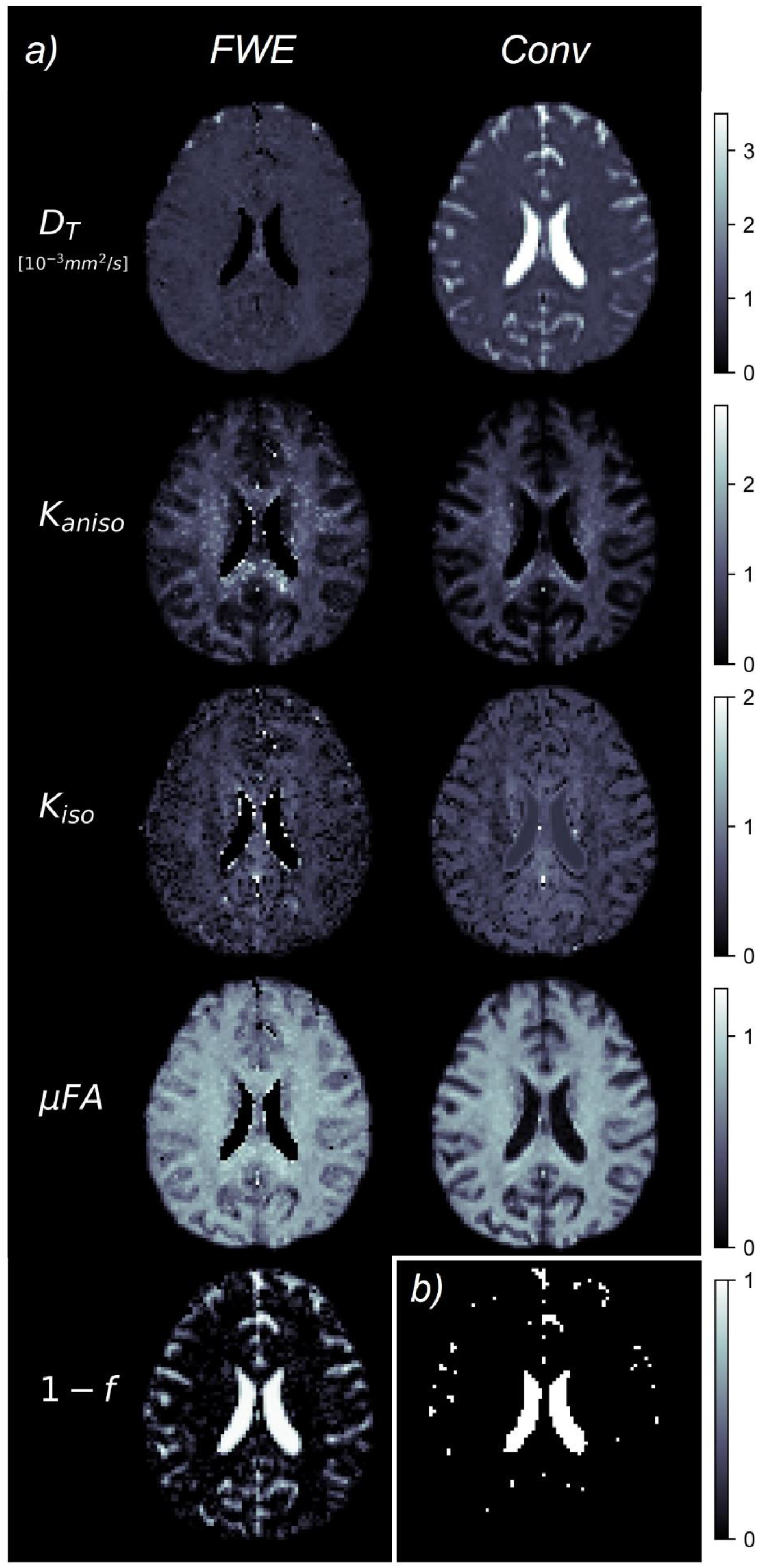
(a) Example slices of tissue diffusivity (*D*_*T*_), anisotropic kurtosis (*K*_*aniso*_), isotropic kurtosis (*K*_*iso*_), microscopic fractional anisotropy (μFA), and fluid volume fraction (1-*f*) measured in one of the healthy volunteers. The images on the left were computed using the free water elimination (FWE) method while those on the right were computed using the conventional (Conv) method. Note that *D*_*T*_ is used interchangeably with *D* for the conventional method. (b) Sample slice depicting a binary map showing voxels with tissue volume fractions less than 0.25, which were omitted in region-of-interest analyses.

**Figure 4.**
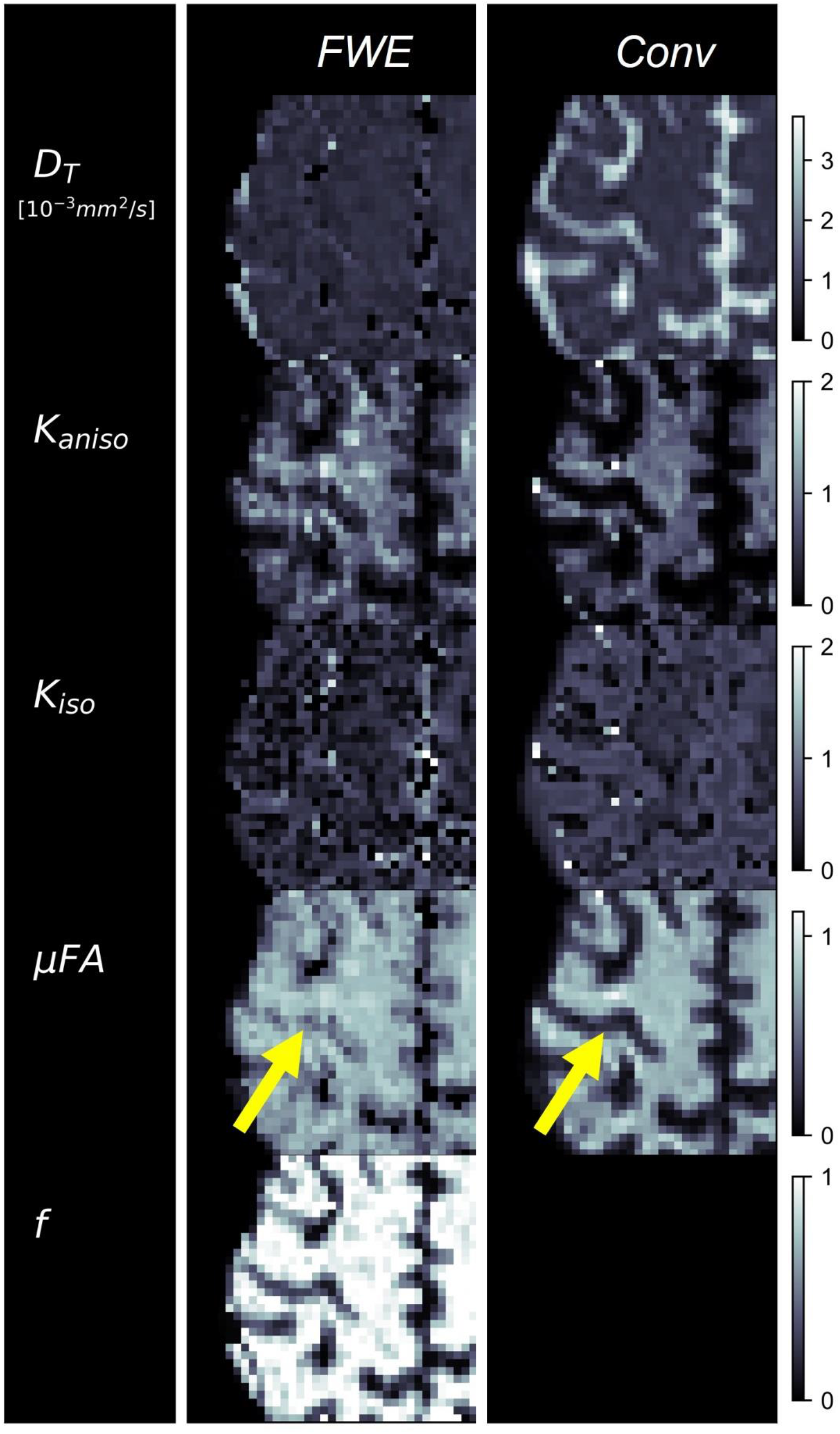
Example cerebral cortex images of tissue diffusivity (*D*_*T*_), anisotropic kurtosis (*K*_*aniso*_), isotropic kurtosis (*K*_*iso*_), microscopic fractional anisotropy (μFA), and tissue volume fraction (*f*) measured in one of the healthy volunteers. The images on the left were computed using the free water elimination (FWE) method while those on the right were computed using the conventional (Conv) method. Note that *D*_*T*_ is used interchangeably with *D* for the conventional method. The yellow arrow highlights a region in which a significant difference is observed between the FWE and conventional μFA measurements due to high free water contamination.

**Figure 5.**
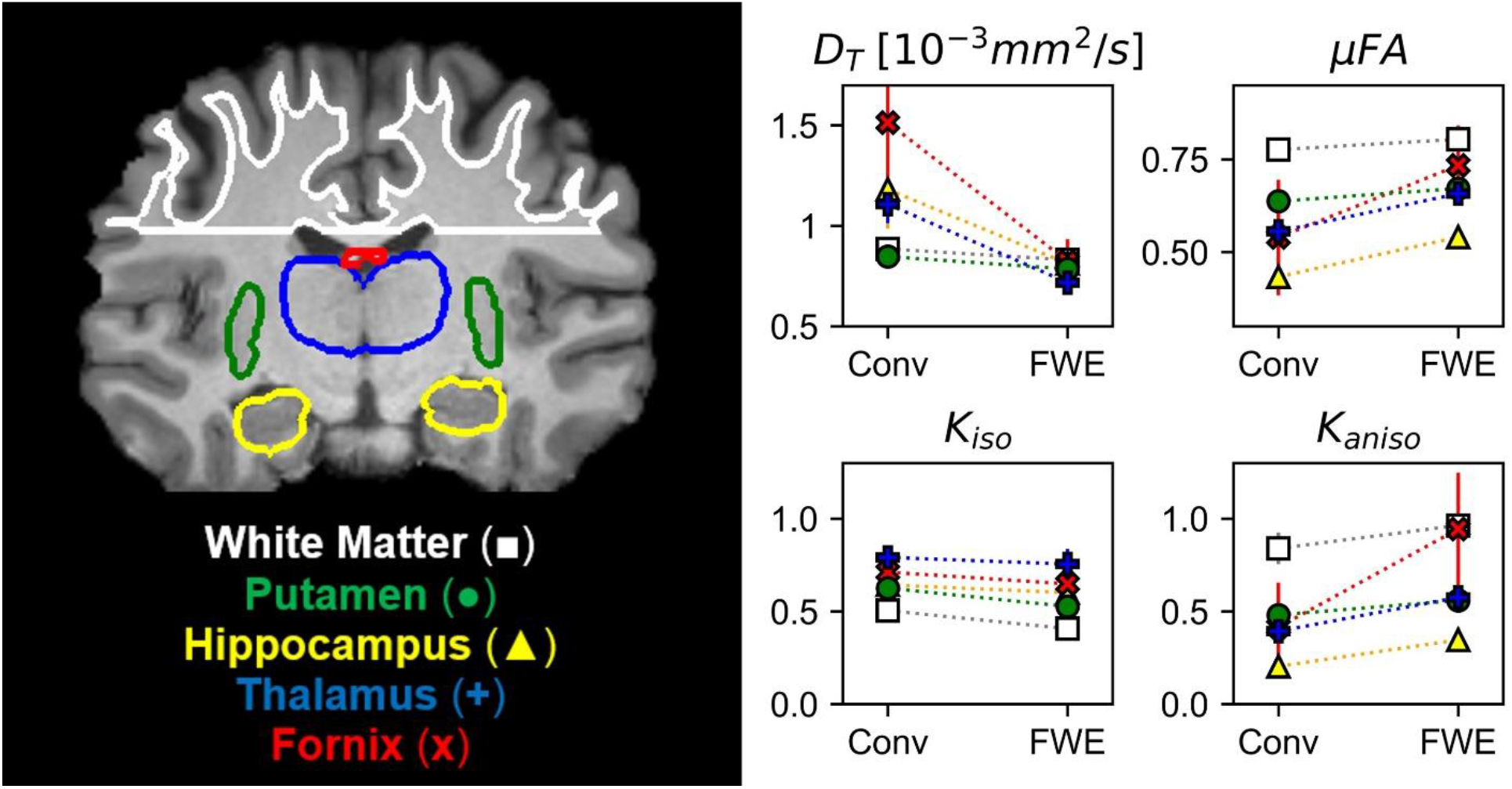
Comparison between the conventional (Conv) and free water elimination (FWE) methods in four healthy volunteers. Depicted on the left is a coronal T1-weighted MPRAGE slice from one of the volunteers highlighting the five regions-of-interest (ROIs). Note that volumetric ROIs were used, despite the single slice depiction. On the right are plots comparing the mean diffusivity (*D*_*T*_), microscopic fractional anisotropy (μFA), isotropic kurtosis (*K*_*iso*_), and anisotropic kurtosis (*K*_*aniso*_) produced by the Conv and FWE methods. In all ROIs, *D*_*T*_ and *K*_*iso*_ were reduced when FWE was applied, while μFA and *K*_*aniso*_ were elevated, though the magnitude of this difference varied by region. Note that *D*_*T*_ is used interchangeably with *D* for the conventional method.

## Discussion

The FWE method presented herein allows for rapid computation of free water-corrected μFA because it uses alternating least squares estimations for *f* and the various diffusion parameters, which are individually rapid. The total processing time was <1 min for each subject. In this work, 100 iterations were performed for each step, but computation time could be further reduced by setting termination criteria for instances where 100 iterations would be excessive. One such example would be to use the estimate of *f* from Part 1 to omit voxels with very high CSF contamination (e.g. *f* < 0.25) from Part 2.

In simulations, the FWE method produced more accurate measurements of *D*_*T*_ and μFA than the conventional method across all volume fractions except *f*=1. At *f*=1, the simulated signal vs. b-value curve has no free water component and resembles the DKI equation (equation 6), so the two-compartment model is redundant and falsely detects a small free water compartment due to the added noise. In simulations with no added noise (data not shown), the FWE and conventional methods both correctly measure *D*_*T*_ and μFA at *f*=1, though only the FWE method yields correct indices at lower volume fractions.

The increase in *K*_*aniso*_ when the FWE method was employed can be explained by the fact that *K*_*aniso*_ arises solely from the tissue compartment. *K*_*aniso*_ describes the variance in the dMRI powder average signal due to the eccentric shape of neuron fibers and other compartments (Jones, Knösche, and Turner 2013); for example, a dMRI acquisition in the direction parallel to neuronal axons will yield a lower signal than one perpendicular to the axons. By removing the isotropic free water compartment, the effect of *K*_*aniso*_ on the remaining signal component is amplified. The reduction in *K*_*iso*_ when the FWE method was used can be attributed to the fact that *K*_*iso*_ describes the variance in diffusivity across compartments; thus, removing the free water compartment, which contains a significantly higher mean diffusivity than neural tissue, attenuates this metric.

While the results of the simulations are promising, it should be noted that the signals were generated using the same free water diffusivity (3*e* − 3 *mm*^2^*/s*) as was assumed in the FWE model, potentially glamorizing the fitting method. In real tissue, deviations from the assumed temperature of 37°C and biases due to differences in T1 and T2 can alter the free water diffusivity and affect the accuracy of the signal fitting algorithm (Pasternak et al. 2009; Pasternak, Maier-Hein, et al. 2014). However, this limitation is shared by all multi-compartment models that use fixed estimates of free water diffusivity and can only be overcome by determining the value prior to the fitting or by attempting to solve for the free water diffusivity in each voxel as an additional variable at the expense of computation time and potential misestimation.

The FWE method also showed promising results when used to measure *D*_*T*_ and μFA in healthy volunteers as differences between the two methods in the various ROIs agreed with expectations. In all ROIs, *D*_*T*_ was reduced and μFA was elevated when the FWE method was used (Fig. 5); these changes are intuitive as removing an isotropic signal compartment with high diffusivity from the overall signal, which also contains anisotropic signal components from neurites and other eccentric compartments, will raise the measured diffusion anisotropy and lower the mean diffusivity. The results of the *in vivo* imaging analysis agreed with the hypotheses that the effects would be smallest in the WM and putamen regions and greatest in the free water-adjacent fornix ROI (Fig. 5). Furthermore, the *f* parameter allowed for the removal of voxels with high CSF contamination from the ROI analysis, improving mean measurements. However, one limitation of the technique is that there are no ground truth measurements to validate the measured indices against. Comparing measured tissue volume fractions against known values from the literature can act as a pseudo-validation of the FWE method, though it should be noted that the measured *f* index represents the T2-weighted signal fraction of the tissue compartment rather than the true volume fraction. To convert *f* to the true volume fraction of tissue, *f*_*T*_, a correction can be made as per the following equation (Veraart, Novikov, and Fieremans 2018):

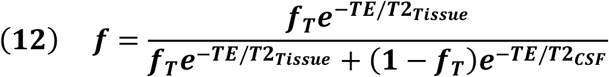

Literature reports free water volume fractions of <2% for WM and 7-9% for GM with high standard deviations (Bender and Klose 2009; Ernst, Kreis, and Ross 1993), which correspond to

*f*_*T*_ values of >0.98 for WM and 0.91-0.93 for GM. Assuming *T*2_*CSF*_ = 1250 ms (Piechnik et al. 2009), *T*2_*WM*_ = 70 ms (Stanisz et al. 2005), and *T*2_*Putamen*_ = *T*2_*Hippocampus*_ = *T*2_*Thalamus*_ = 95 ms (Stanisz et al. 2005; Bartlett et al. 2007) at 3T, the approximate mean *f*_*T*_ values for the WM, putamen, hippocampus, and thalamus regions were 0.99, 0.98, 0.92, and 0.92 in the healthy volunteers imaged in this work. As expected, the volume fraction in the fornix was measured to be much lower than the other ROIs (*f* = 0.64); no correction was performed for this region because many voxels contained large volumes of pure CSF, which violates the assumptions of equation 12.

There are several limitations potentially affecting this study. Diffusion time discrepancies between the LTE and STE sequences, and between the three gradient channels in the STE sequence, were not taken into consideration in this work. Different diffusion times in the LTE and STE acquisitions could lead to slight differences between the respective powder average signals that are misattributed to be differences between *K*_*LTE*_ and *K*_*STE*_, while different diffusion times in the different gradient channels for the STE acquisitions could give rise to orientational biases (Jespersen et al. 2019). These potential biases are not expected to have had a significant effect on the results of this work since both the FWE method and conventional method were applied to the same data, and any biases caused by time-dependent diffusion would affect both models. However, future studies should consider using optimized STE sequences to ensure that the diffusion time of the STE and LTE sequences match and that there are no orientational biases in the STE sequence.

Another limitation is that the time required for an acquisition protocol to acquire powder average signals at 4 b-values in both LTE and STE schema, could be prohibitively long for some applications (our *in vivo* scan required 9 min).

Both the kurtosis and FWE models used herein assume that any deviation from mono-Gaussian diffusion in the powder average dMRI signals arises exclusively from two distinct sources: *K*_*aniso*_ and *K*_*iso*_. However, restricted diffusion inside compartments, exchange between compartments, and microstructural disorder can also contribute to the overall kurtosis and are often categorized together in a term known as microscopic kurtosis (*K*_µ_) (Jespersen et al. 2019; Henriques, Jespersen, and Shemesh 2020). Though most μFA imaging techniques do not consider

*K*_µ_, recent studies have found that it is non-negligible in the human brain and that ignoring it can lead to biases (Novello et al. 2022). Despite this limitation, μFA techniques that don’t distinguish *K*_µ_ from other kurtosis sources have shown promising diagnostic and research capabilities and still represent a significant advance over the widely used DTI metrics.

The images produced by the FWE method (Fig. 3) appear grainier than those produced by the conventional method and higher standard deviations were measured in all metrics when the FWE method was used, both in simulations (Figs. 1 and 2) and *in vivo* (Fig. 5). This increased variance is expected due to the increased complexity of the FWE-DKI model relative to the DKI model. Studies that use the FWE technique should design MRI protocols that sample more b-shells to improve the data fit and acquire more LTE and STE scans at each b-value to raise the SNR of the powder average signals. A minimalistic protocol, such as those described in the literature (Nilsson et al. 2019; Arezza, Tse, and Baron 2021), may be insufficient for FWE imaging. Also, regularization enforcing spatial smoothness, similar to that applied in other applications of FWE, could likely help mitigate this issue (Golub, Neto Henriques, and Gouveia Nunes 2020; Pasternak et al. 2009).

In conclusion, the two-compartment μFA imaging technique presented in this work represents an extension to the μFA imaging technique that integrates a free water compartment to extract tissue-specific indices of *D, K*_*aniso*_, *K*_*iso*_, and μFA. To solve the ill-conditioned fit of the data to equations (8) and (9), a two-part algorithm was employed to first determine initial guesses of key parameters and then to perform the joint fit. Any dMRI protocol designed to estimate μFA via the FWE method will be versatile due to the need for multiple b-shells in both LTE and STE schema and can also be fitted to the conventional μFA method and the DKI model; furthermore, if a significant number of LTE directions are acquired at b = 1000 s/mm^2^, the data can be fitted to the widely adopted DTI model as well. Both simulation and real data experiments indicated that the FWE method may be a feasible technique for measuring μFA and other dMRI indices with greater specificity to neural tissue characteristics by removing free water partial volume effects.

## Data Availability

All data produced are available online at https://osf.io/etkgx/

## Author Contributions

NA: conceptualization, methodology, experiments, and writing. MO: preliminary experiments. CB: conceptualization, methodology, review, and supervision.

## References

Alexander, A. L., K. M. Hasan, M. Lazar, J. S. Tsuruda, and D. L. Parker. 2001. “Analysis of Partial Volume Effects in Diffusion-Tensor MRI.” Magnetic Resonance in Medicine: Official Journal of the Society of Magnetic Resonance in Medicine / Society of Magnetic Resonance in Medicine 45 (5): 770–80.

Andersson, Jesper L. R., and Stamatios N. Sotiropoulos. 2016. “An Integrated Approach to Correction for Off-Resonance Effects and Subject Movement in Diffusion MR Imaging.” NeuroImage 125 (January): 1063–78.

Arezza, N. J. J., D. H. Y. Tse, and C. A. Baron. 2021. “Rapid Microscopic Fractional Anisotropy Imaging via an Optimized Linear Regression Formulation.” Magnetic Resonance Imaging 80 (May): 132–43.

Baron, Corey A., and Christian Beaulieu. 2015. “Acquisition Strategy to Reduce Cerebrospinal Fluid Partial Volume Effects for Improved DTI Tractography.” Magnetic Resonance in Medicine: Official Journal of the Society of Magnetic Resonance in Medicine / Society of Magnetic Resonance in Medicine 73 (3): 1075–84.

Baron, Corey, and Nico Joseph John Arezza. 2020. “Test-Retest Data Repository for Spherical Tensor Encoding.” OSF. October 14, 2020. https://osf.io/etkgx/.

Bartlett, P. A., M. R. Symms, S. L. Free, and J. S. Duncan. 2007. “T2 Relaxometry of the Hippocampus at 3T.” AJNR. American Journal of Neuroradiology 28 (6): 1095–98.

Basser, P. J., J. Mattiello, and D. LeBihan. 1994. “MR Diffusion Tensor Spectroscopy and Imaging.” Biophysical Journal 66 (1): 259–67.

Bender, Benjamin, and Uwe Klose. 2009. “Cerebrospinal Fluid and Interstitial Fluid Volume Measurements in the Human Brain at 3T with EPI.” Magnetic Resonance in Medicine: Official Journal of the Society of Magnetic Resonance in Medicine / Society of Magnetic Resonance in Medicine 61 (4): 834–41.

Chou, Ming-Chung, Yi-Ru Lin, Teng-Yi Huang, Chao-Ying Wang, Hsiao-Wen Chung, Chun-Jung Juan, and Cheng-Yu Chen. 2005. “FLAIR Diffusion-Tensor MR Tractography: Comparison of Fiber Tracking with Conventional Imaging.” AJNR. American Journal of Neuroradiology 26 (3): 591–97.

Collier, Quinten, Jelle Veraart, Ben Jeurissen, Floris Vanhevel, Pim Pullens, Paul M. Parizel, Arnold J. den Dekker, and Jan Sijbers. 2018. “Diffusion Kurtosis Imaging with Free Water Elimination: A Bayesian Estimation Approach.” Magnetic Resonance in Medicine: Official Journal of the Society of Magnetic Resonance in Medicine / Society of Magnetic Resonance in Medicine 80 (2): 802–13.

Cory, D. G., A. N. Garroway, and J. B. Miller. 1990. “Applications of Spin Transport as a Probe of Local Geometry.” In Abstracts of Papers of the American Chemical Society, 199:105-POLY. AMER CHEMICAL SOC 1155 16TH ST, NW, WASHINGTON, DC 20036.

Ernst, T., R. Kreis, and B. D. Ross. 1993. “Absolute Quantitation of Water and Metabolites in the Human Brain. I. Compartments and Water.” Journal of Magnetic Resonance. Series B 102 (1): 1–8.

Everdingen, K. J. van, J. van der Grond, L. J. Kappelle, L. M. Ramos, and W. P. Mali. 1998. “Diffusion-Weighted Magnetic Resonance Imaging in Acute Stroke.” Stroke; a Journal of Cerebral Circulation 29 (9): 1783–90.

Frisken, B. J. 2001. “Revisiting the Method of Cumulants for the Analysis of Dynamic Light-Scattering Data.” Applied Optics 40 (24): 4087–91.

Golub, Marc, Rafael Neto Henriques, and Rita Gouveia Nunes. 2020. “Free-Water DTI Estimates from Single b-Value Data Might Seem Plausible but Must Be Interpreted with Care.” Magnetic Resonance in Medicine: Official Journal of the Society of Magnetic Resonance in Medicine / Society of Magnetic Resonance in Medicine, December. https://doi.org/10.1002/mrm.28599.

Henriques, Rafael Neto, Sune Nørhøj Jespersen, and Noam Shemesh. 2020. “Correlation Tensor Magnetic Resonance Imaging.” NeuroImage 211 (May): 116605.

Hoy, Andrew R., Martina Ly, Cynthia M. Carlsson, Ozioma C. Okonkwo, Henrik Zetterberg, Kaj Blennow, Mark A. Sager, et al. 2017. “Microstructural White Matter Alterations in Preclinical Alzheimer’s Disease Detected Using Free Water Elimination Diffusion Tensor Imaging.” PloS One 12 (3): e0173982.

Ikenouchi, Yutaka, Koji Kamagata, Christina Andica, Taku Hatano, Takashi Ogawa, Haruka Takeshige-Amano, Kouhei Kamiya, et al. 2020. “Evaluation of White Matter Microstructure in Patients with Parkinson’s Disease Using Microscopic Fractional Anisotropy.” Neuroradiology 62 (2): 197–203.

Inglese, M., and Maxim Bester. 2010. “Diffusion Imaging in Multiple Sclerosis: Research and Clinical Implications.” NMR in Biomedicine. https://doi.org/10.1002/nbm.1515.

Jensen, Jens H., Joseph A. Helpern, Anita Ramani, Hanzhang Lu, and Kyle Kaczynski. 2005. “Diffusional Kurtosis Imaging: The Quantification of Non-Gaussian Water Diffusion by Means of Magnetic Resonance Imaging.” Magnetic Resonance in Medicine: An Official Journal of the International Society for Magnetic Resonance in Medicine 53 (6): 1432–40.

Jespersen, Sune Nørhøj, Henrik Lundell, Casper Kaae Sønderby, and Tim B. Dyrby. 2013. “Orientationally Invariant Metrics of Apparent Compartment Eccentricity from Double Pulsed Field Gradient Diffusion Experiments.” NMR in Biomedicine 26 (12): 1647–62.

Jespersen, Sune Nørhøj, Jonas Lynge Olesen, Andrada Ianuş, and Noam Shemesh. 2019. “Effects of Nongaussian Diffusion on ‘Isotropic Diffusion’ Measurements: An Ex-Vivo Microimaging and Simulation Study.” Journal of Magnetic Resonance 300 (March): 84–94.

Johansen-Berg, Heidi, and Timothy E. J. Behrens. 2009. Diffusion MRI: From Quantitative Measurement to In-Vivo Neuroanatomy. Vol. 1. Elsevier/Academic Press.

Jones, Derek K., and Mara Cercignani. 2010. “Twenty-Five Pitfalls in the Analysis of Diffusion MRI Data.” NMR in Biomedicine 23 (7): 803–20.

Jones, Derek K., Thomas R. Knösche, and Robert Turner. 2013. “White Matter Integrity, Fiber Count, and Other Fallacies: The Do’s and Don’ts of Diffusion MRI.” NeuroImage 73 (June): 239–54.

Kaden, Enrico, Nathaniel D. Kelm, Robert P. Carson, Mark D. Does, and Daniel C. Alexander. 2016. “Multi-Compartment Microscopic Diffusion Imaging.” NeuroImage 139 (October): 346–59.

Kaden, Enrico, Frithjof Kruggel, and Daniel C. Alexander. 2016. “Quantitative Mapping of the Per-Axon Diffusion Coefficients in Brain White Matter.” Magnetic Resonance in Medicine: Official Journal of the Society of Magnetic Resonance in Medicine / Society of Magnetic Resonance in Medicine 75 (4): 1752–63.

Kellner, Elias, Bibek Dhital, Valerij G. Kiselev, and Marco Reisert. 2016. “Gibbs-Ringing Artifact Removal Based on Local Subvoxel-Shifts.” Magnetic Resonance in Medicine: Official Journal of the Society of Magnetic Resonance in Medicine / Society of Magnetic Resonance in Medicine 76 (5): 1574–81.

Lasič, Samo, Filip Szczepankiewicz, Stefanie Eriksson, Markus Nilsson, and Daniel Topgaard. 2014. “Microanisotropy Imaging: Quantification of Microscopic Diffusion Anisotropy and Orientational Order Parameter by Diffusion MRI with Magic-Angle Spinning of the q-Vector.” Frontiers in Physics 2. https://doi.org/10.3389/fphy.2014.00011.

Lawrenz, Marco, Stefanie Brassen, and Jürgen Finsterbusch. 2016. “Microscopic Diffusion Anisotropy in the Human Brain: Age-Related Changes.” NeuroImage 141 (November): 313–25.

Li, Shumei, Bin Wang, Pengfei Xu, Qixiang Lin, Gaolang Gong, Xiaoling Peng, Yuanyuan Fan, Yong He, and Ruiwang Huang. 2013. “Increased Global and Local Efficiency of Human Brain Anatomical Networks Detected with FLAIR-DTI Compared to Non-FLAIR-DTI.” PloS One 8 (8): e71229.

Metzler-Baddeley, Claudia, Michael J. O’Sullivan, Sonya Bells, Ofer Pasternak, and Derek K. Jones. 2012. “How and How Not to Correct for CSF-Contamination in Diffusion MRI.” NeuroImage 59 (2): 1394–1403.

Nilsson, Markus, Filip Szczepankiewicz, Jan Brabec, Marie Taylor, Carl-fredrik Westin, Alexandra Golby, Danielle Westen, and Pia C. Sundgren. 2019. “Tensor-valued Diffusion MRI in under 3 Minutes: An Initial Survey of Microscopic Anisotropy and Tissue Heterogeneity in Intracranial Tumors.” Magnetic Resonance in Medicine: Official Journal of the Society of Magnetic Resonance in Medicine / Society of Magnetic Resonance in Medicine, September. https://doi.org/10.1002/mrm.27959.

Novello, Lisa, Rafael Neto Henriques, Andrada Ianuş, Thorsten Feiweier, Noam Shemesh, and Jorge Jovicich. 2022. “In Vivo Correlation Tensor MRI Reveals Microscopic Kurtosis in the Human Brain on a Clinical 3T Scanner.” NeuroImage 254 (July): 119137.

Novikov, Dmitry S., Els Fieremans, Sune N. Jespersen, and Valerij G. Kiselev. 2019. “Quantifying Brain Microstructure with Diffusion MRI: Theory and Parameter Estimation.” NMR in Biomedicine 32 (4): e3998.

Papadakis, Nikos G., Kay M. Martin, Mohammed H. Mustafa, Iain D. Wilkinson, Paul D. Griffiths, Chris L-H Huang, and Peter W. R. Woodruff. 2002. “Study of the Effect of CSF Suppression on White Matter Diffusion Anisotropy Mapping of Healthy Human Brain.” Magnetic Resonance in Medicine: Official Journal of the Society of Magnetic Resonance in Medicine / Society of Magnetic Resonance in Medicine 48 (2): 394–98.

Pasternak, Ofer, Inga K. Koerte, Sylvain Bouix, Eli Fredman, Takeshi Sasaki, Michael Mayinger, Karl G. Helmer, et al. 2014. “Hockey Concussion Education Project, Part 2. Microstructural White Matter Alterations in Acutely Concussed Ice Hockey Players: A Longitudinal Free-Water MRI Study.” Journal of Neurosurgery 120 (4): 873–81.

Pasternak, Ofer, Klaus Maier-Hein, Christian Baumgartner, Martha E. Shenton, Yogesh Rathi, and Carl-Fredrik Westin. 2014. “The Estimation of Free-Water Corrected Diffusion Tensors.” In Visualization and Processing of Tensors and Higher Order Descriptors for Multi-Valued Data, 249–70. Springer Berlin Heidelberg.

Pasternak, Ofer, Nir Sochen, Yaniv Gur, Nathan Intrator, and Yaniv Assaf. 2009. “Free Water Elimination and Mapping from Diffusion MRI.” Magnetic Resonance in Medicine: Official Journal of the Society of Magnetic Resonance in Medicine / Society of Magnetic Resonance in Medicine 62 (3): 717–30.

Patenaude, Brian, Stephen M. Smith, David N. Kennedy, and Mark Jenkinson. 2011. “A Bayesian Model of Shape and Appearance for Subcortical Brain Segmentation.” NeuroImage 56 (3): 907–22.

Piechnik, S. K., J. Evans, L. H. Bary, R. G. Wise, and P. Jezzard. 2009. “Functional Changes in CSF Volume Estimated Using Measurement of Water T2 Relaxation.” Magnetic Resonance in Medicine: Official Journal of the Society of Magnetic Resonance in Medicine / Society of Magnetic Resonance in Medicine 61 (3): 579–86.

Pierpaoli, C., and P. J. Basser. 1996. “Toward a Quantitative Assessment of Diffusion Anisotropy.” Magnetic Resonance in Medicine: Official Journal of the Society of Magnetic Resonance in Medicine / Society of Magnetic Resonance in Medicine 36 (6): 893–906.

Planetta, Peggy J., Edward Ofori, Ofer Pasternak, Roxana G. Burciu, Priyank Shukla, Jesse C. DeSimone, Michael S. Okun, Nikolaus R. McFarland, and David E. Vaillancourt. 2016. “Free-Water Imaging in Parkinson’s Disease and Atypical Parkinsonism.” Brain: A Journal of Neurology 139 (Pt 2): 495–508.

Rovaris, M., A. Gass, R. Bammer, S. J. Hickman, O. Ciccarelli, D. H. Miller, and M. Filippi. 2005. “Diffusion MRI in Multiple Sclerosis.” Neurology 65 (10): 1526–32.

Shemesh, Noam, Sune N. Jespersen, Daniel C. Alexander, Yoram Cohen, Ivana Drobnjak, Tim B. Dyrby, Jurgen Finsterbusch, et al. 2016. “Conventions and Nomenclature for Double Diffusion Encoding NMR and MRI.” Magnetic Resonance in Medicine: Official Journal of the Society of Magnetic Resonance in Medicine / Society of Magnetic Resonance in Medicine 75 (1): 82–87.

Stanisz, Greg J., Ewa E. Odrobina, Joseph Pun, Michael Escaravage, Simon J. Graham, Michael J. Bronskill, and R. Mark Henkelman. 2005. “T1, T2 Relaxation and Magnetization Transfer in Tissue at 3T.” Magnetic Resonance in Medicine: Official Journal of the Society of Magnetic Resonance in Medicine / Society of Magnetic Resonance in Medicine 54 (3): 507–12.

Stejskal, E. O., and J. E. Tanner. 1965. “Spin Diffusion Measurements: Spin Echoes in the Presence of a Time-Dependent Field Gradient.” The Journal of Chemical Physics 42 (1): 288–92.

Szczepankiewicz, Filip, Samo Lasič, Danielle van Westen, Pia C. Sundgren, Elisabet Englund, Carl Fredrik Westin, Freddy Ståhlberg, Jimmy Lätt, Daniel Topgaard, and Markus Nilsson. 2015. “Quantification of Microscopic Diffusion Anisotropy Disentangles Effects of Orientation Dispersion from Microstructure: Applications in Healthy Volunteers and in Brain Tumors.” NeuroImage 104 (January): 241–52.

Tanner, J. E. 1965. “Pulsed Field Gradients for NMR Spin-Echo Diffusion Measurements.” The Review of Scientific Instruments 36 (8): 1086–87.

Tournier, J-Donald, Robert Smith, David Raffelt, Rami Tabbara, Thijs Dhollander, Maximilian Pietsch, Daan Christiaens, Ben Jeurissen, Chun-Hung Yeh, and Alan Connelly. 2019. “MRtrix3: A Fast, Flexible and Open Software Framework for Medical Image Processing and Visualisation.” NeuroImage 202 (November): 116137.

Veraart, Jelle, Dmitry S. Novikov, Daan Christiaens, Benjamin Ades-Aron, Jan Sijbers, and Els Fieremans. 2016. “Denoising of Diffusion MRI Using Random Matrix Theory.” NeuroImage 142 (November): 394–406.

Veraart, Jelle, Dmitry S. Novikov, and Els Fieremans. 2018. “TE Dependent Diffusion Imaging (TEdDI) Distinguishes between Compartmental T2 Relaxation Times.” NeuroImage 182 (November): 360–69.

Vos, Sjoerd B., Derek K. Jones, Max A. Viergever, and Alexander Leemans. 2011. “Partial Volume Effect as a Hidden Covariate in DTI Analyses.” NeuroImage 55 (4): 1566–76.

Westin, Carl-Fredrik, Hans Knutsson, Ofer Pasternak, Filip Szczepankiewicz, Evren Özarslan, Danielle van Westen, Cecilia Mattisson, et al. 2016. “Q-Space Trajectory Imaging for Multidimensional Diffusion MRI of the Human Brain.” NeuroImage 135 (July): 345–62.

Yang, Grant, Qiyuan Tian, Christoph Leuze, Max Wintermark, and Jennifer A. McNab. 2018. “Double Diffusion Encoding MRI for the Clinic.” Magnetic Resonance in Medicine: Official Journal of the Society of Magnetic Resonance in Medicine / Society of Magnetic Resonance in Medicine 80 (2): 507–20.

Yoo, Jiyoon, Leevi Kerkelä, Patrick W. Hales, Kiran K. Seunarine, and Christopher A. Clark. 2021. “High-Resolution Microscopic Diffusion Anisotropy Imaging in the Human Hippocampus at 3T.” Magnetic Resonance in Medicine: Official Journal of the Society of Magnetic Resonance in Medicine / Society of Magnetic Resonance in Medicine, November. https://doi.org/10.1002/mrm.29104.

Zhang, Y., M. Brady, and S. Smith. 2001. “Segmentation of Brain MR Images through a Hidden Markov Random Field Model and the Expectation-Maximization Algorithm.” IEEE Transactions on Medical Imaging 20 (1): 45–57.

Zhang, Yu, Norbert Schuff, An-Tao Du, Howard J. Rosen, Joel H. Kramer, Maria Luisa Gorno-Tempini, Bruce L. Miller, and Michael W. Weiner. 2009. “White Matter Damage in Frontotemporal Dementia and Alzheimer’s Disease Measured by Diffusion MRI.” Brain: A Journal of Neurology 132 (Pt 9): 2579–92.

